# Clinical manifestations of hospitalized COVID-19 patients in Bangladesh: a 14-day observational study

**DOI:** 10.1101/2021.04.27.21256018

**Authors:** Modhusudon Shaha, Md. Azizul Islam, Faizul Huq, Bithi Roy, Ashraful Kabir, Md. Salimullah, Mamun-Al-Mahtab, Sheikh Mohammad Fazle Akbar

## Abstract

**Objectives:** Severe acute respiratory coronavirus-2 (SARS-CoV-2) is currently a significant public health concern and causing a pandemic in the world. Despite of immense attention to the coronavirus disease-2019 (COVID-19), very little attention has been given to the kinetics of disease progression in infected patients. Therefore, in this study, we present a 14-day clinical observation of hospital-admitted COVID-19 patients.

**Methods:** After recording the demography of 42 COVID-19 patients on day 1, we observed the clinical progression for 14 days by investigating the hematological and biochemical responses of patients blood and serum, respectively.

**Results:** Approximately 62% of the hospital-admitted COVID-19 patients presented cough, followed by fever (approximately 52%). The top comorbidities of these patients were hypertension (30%) and diabetes mellitus (19%). The average blood hemoglobin level was slightly low among the patients in early days of infection and went up to the normal level on the latter days. A substantial increase in the level of ALT/SGPT (up to 106 units/L; SEM: 12.64) and AST/SGOT (up to 64.35 units/L; SEM: 5.013) in COVID-19 patients was observed, which may suggest that infection with CoV-2019 is associated with the functionality of other organs of COVID-19 patients.

**Conclusion:** This 14-day observational study may help clinicians to decide the choice of treatment of COVID-19 patients.

## Introduction

Coronaviruses have been causing significant disease outbreaks globally over the past two decades. In 2002, severe acute respiratory syndrome (SARS) emerged in East Asia (Cherry 2004), and later on, the Middle East respiratory syndrome (MERS) was associated with an outbreak in the Middle East in 2012 (Milne-Price, Miazgowicz, and Munster 2014). Recently, a novel coronavirus, SARS-coronavirus-2019 (SARS-CoV-2), have emerged in late 2019 in China and posed a threat to global health, causing coronavirus disease 2019 (COVID-19) (Rodriguez-Morales et al. 2020). This virus is currently causing a pandemic in the world. The novel SARS-CoV-2 was first reported in Wuhan, China on December 12, 2019 (Zhu et al. 2020). Globally, health workers are currently making tremendous attempts to control the ongoing disease outbreaks caused by this virus.

Newly emerged SARS-CoV-2 poses a threat to the current public health worldwide. The source of infection of this third coronavirus outbreak in humans is not identified yet. After appearing in Wuhan, this virus has spread rapidly throughout the country and, eventually, to other countries. Due to the rapidly spreading nature of this virus on a global scale and the severity of the ongoing outbreak, the WHO announced a Public Health Emergency of International Concern (PHEIC) on January 31, 2020 (WHO 2020). Later on March 11, 2020, they declared the situation a pandemic. At the latest, new variants of this virus have emerged in some countries, including Bangladesh (Kirby 2021; Le Page 2021; M.E. Hossain, Rahman, et al. 2021; M.M. Hasan, Das, et al. 2021; Moniruzzaman et al. 2020; M.U. Hossain, Bhattacharjee, et al. 2021). After developing infection in the host, the virus exploits the immune system of the respective host in a very rapid time and poses threats to the hosts’ life. However, the clinical manifestations of how the infection progresses to the severe disease and/or subsequently recovers are yet to be understood. Here in this study, we present the kinetics of clinical manifestations of the hospital-admitted COVID-19 patients for 14 days and patients’ comorbidity with the COVID-19 disease.

## Materials and Methods

### Selection of patients and sample collection

The study was conducted between April and May 2020 at Combined Military Hospital, Dhaka, Bangladesh. Forty-two patients were included in this study who tested COVID-19 positive and were admitted to the hospital. The mean age of these patients was 40.02 years, with a standard error of the mean (SEM) of 1.911. Of these patients, female (mean age: 32.13; SEM: 5.313) and male (mean age: 41.88; SEM: 1.91) patients were approximately 19% and 81%, respectively. The demographic of these patients were collected on the day of hospital admission. The enrolled patients were in observation for 14 days (unless otherwise stated), and their blood samples were collected each day at the same time during the course of observation. For hematological analyses, blood samples of the COVID-19 patients were collected in tubes containing anticoagulant EDTA. For performing serum biochemistry, blood samples were collected without anticoagulant and was centrifuged at 500 g for 10 min to separate serum (Eppendorf 5702, Hamburg, Germany). The separated serum was then stored at -20□ until use. This study was approved by the institutional ethics committee (approval number: FGH/2020/01) of Farabi General Hospital, Dhaka, Bangladesh.

### Hematological and biochemical analyses

A complete blood count (CBC) using the COVID-19 patients’ blood samples was performed using the commercial kit (Promega corporation, Fitchburg, WI, USA) using ADVIA 2120 Hematology System (Siemens Diagnostics, Erlangen, Germany) according to the manufacturer’s instructions. Blood oxygen was measured using a pulse oximeter (Thermo Fisher Scientific, Waltham, MA, USA) as per the protocol given by the supplier.

Biochemical analyses of serum components such as Albumin, Bilirubin, creatinine, chloride, sodium, and potassium were performed using standard routine techniques. Serum AST and ALT levels were determined by the standard colorimetric method of Reitman and Frankel. Enzyme activities were determined at 37°C. Serum biochemistry was performed using a standard autoanalyzer (ABX-Diagnostics, Tokyo, Japan).

### Other relevant observations and statistical analysis

Body temperature, respiratory rate, blood pressure, and heart rate of the COVID-19 patients were measured as routine procedures in the lab. Chest X-ray reports were kindly provided by the imaging unit of the hospital on day 1. GCS score was determined as per the standard protocol (Teasdale and Jennett 1974). All statistical analysis was performed using GraphPad Prism v9.

## Results

### Differential clinical symptoms and comorbidities were presented by the COVID-19 patients

Of the 42 hospital-admitted COVID-19 patients, approximately 62% were affected by cough, followed by fever (∼52%), breathing difficulty (∼24%), myalgia (∼22%), loose motion (∼17%), and others (**Figure 1A**). Of these patients, approximately 19% got pneumonia, and only about 2% were asymptomatic. These results indicate that in addition to fever and cough, COVID-19-infected patients presented a wide range of other clinical symptoms.

**Figure 1:**
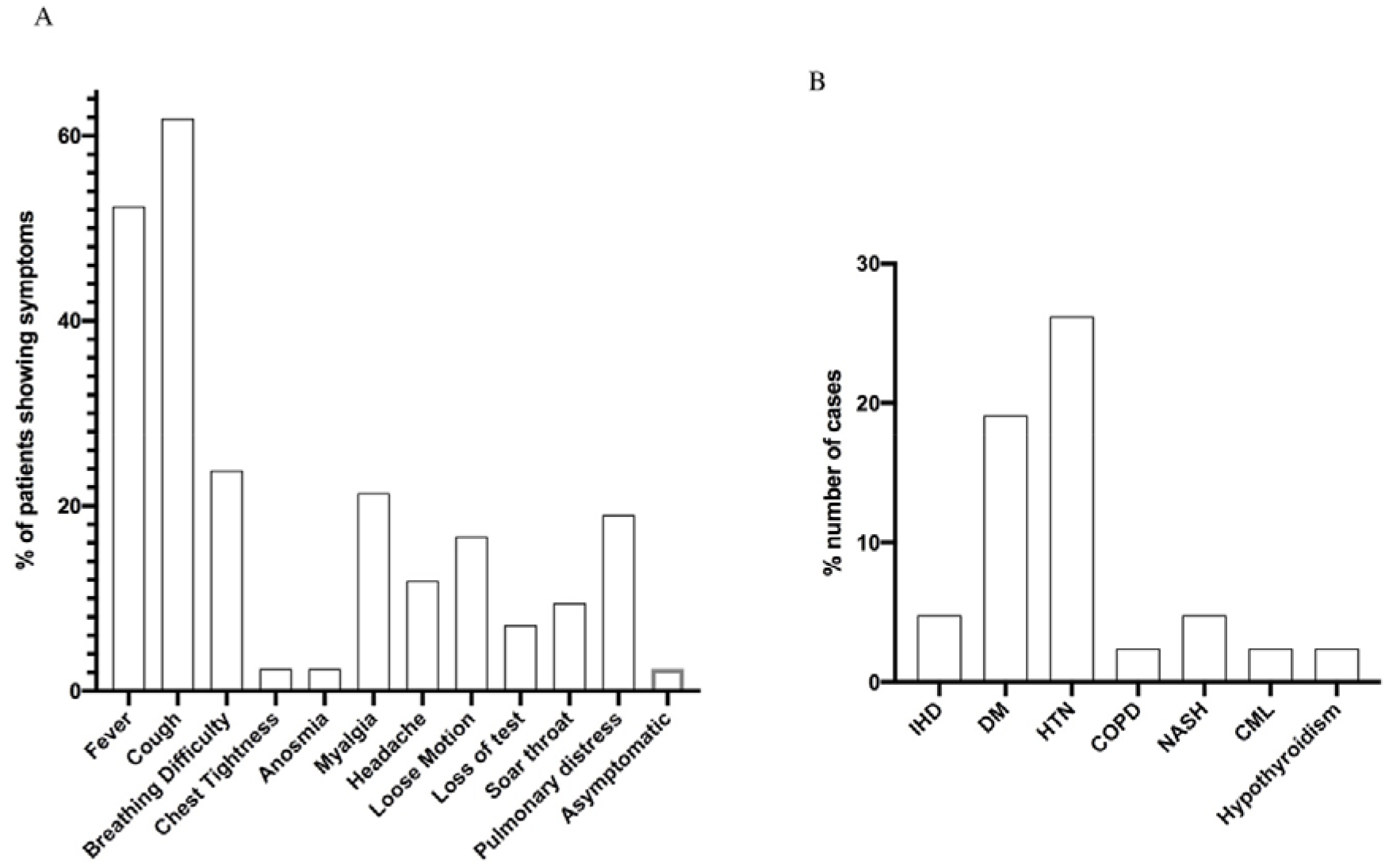
Clinical symptoms (A) and comorbidity (B) presented by the hospital-admitted COVID-19 patients. Bar chart was prepared using GraphPad Prism v9 software package. Data is presented as the percentage (%) of the total included patients. Abbreviations for Fig1B: IHD- Ischemic Heart Disease; DM-Diabetes Mellitus; HTN-Hypertension; COPD-Chronic Obstructive Pulmonary Disease; NASH- Non-alcoholic Steatohepatitis; CML-Chronic Myelogenous Leukemia.

Chronic conditions were reported as common comorbidities among the COVID19 patients. Data revealed that nearly 30% of the included patients were affected with hypertension. Diabetes Mellitus was the second comorbid condition, as reported by approximately 19%. Ischemic Heart Disease (5%), Non-alcoholic Steatohepatitis (6%), Hypothyroidism (<5%) were the other common comorbid condition among the patients (**Figure 1B)**. At that time of COVID-19 pandemic, the patients were treated with Fevipiravir plus SoC (∼19%), Hydroxychloroqine plus azithromycin plus SoC (∼38.1%), and Hydroxychloroqine plus doxycycline plus SoC (54.8%). Approximately 14.3% of the patients was given a combination of Fevipiravir and Hydroxychloroqine plus doxycycline plus SoC therapy at that time. Only 1 (2.4%) patient needed oxygen during the course of observation.

### Increased respiratory rate of COVID-19 patients

As the COVID-19 patients present a wide range of clinical symptoms and comorbidities related to both respiratory and heart organs, we determined the respiratory rate as well as the heart rate of these patients from day 1 to day 14. We observed a higher mean respiratory rate (25.07 breaths/min; SEM: 0.6652; normal range 12-16 breaths/min) among the COVID-19 patients on day 1. The respiratory rate of these patients fluctuated slightly during the course of observation but remained higher than the normal range, as shown in **Figure 2**. However, the heart rate of these patients was in the normal range during the course of observation.

**Figure 2:**
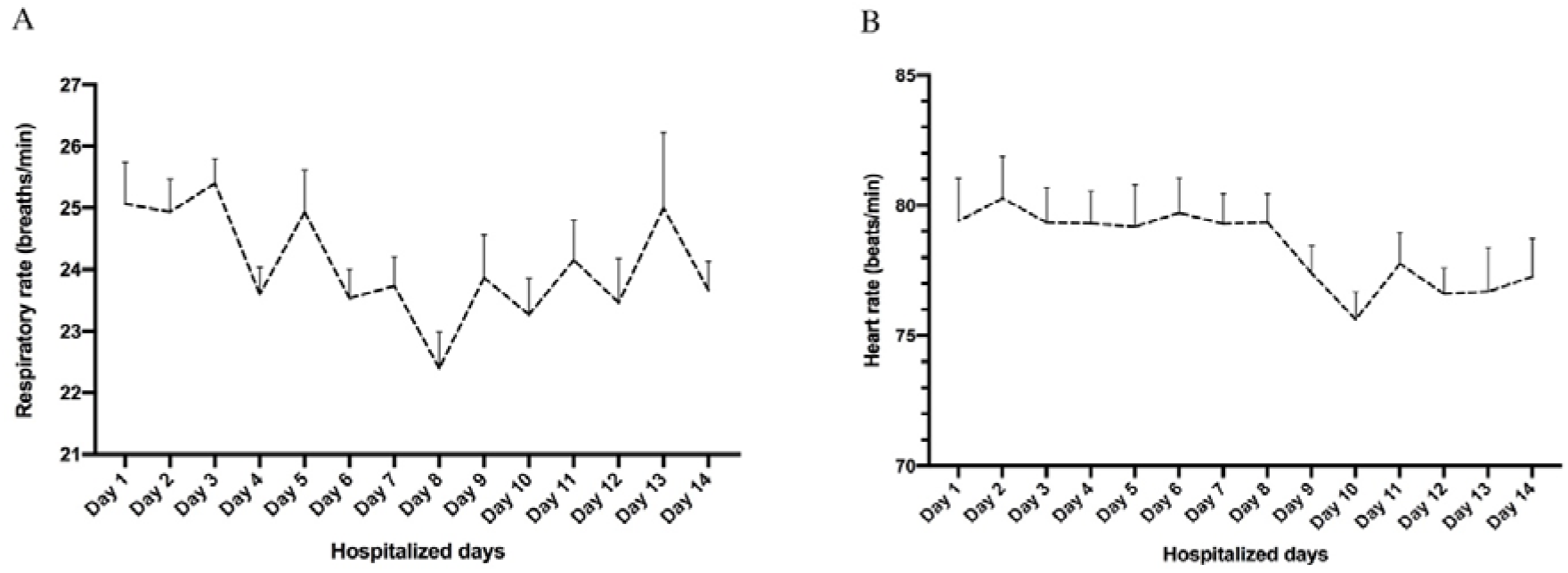
A 14-day observation of respiratory rate (A) and heart rate (B) of the COVID-19 patients. The graph was prepared using GraphPad Prism v9 software package. The represented data denotes Mean±SEM.

We also measured the body temperature of the COVID-19 patients in the course of observation. Analysis of data revealed that the mean body temperature of the patients gradually decreased from the hospital’s 1st day (99.42° F; SEM: 0.3614) to the 11th day (96.71° F; SEM: 0.3555). There was an increase on the 12th day (97.43° F; SEM: 0.321). The mean temperature was recorded on the 14th day as 96.69° F (SEM: 0.4606) (data not shown).

### Irregular level of blood components in COVID-19 patients

After observing a differential level of respiratory rate and body temperature of the COVID-19 patients in different days of hospital stay, we hypothesized that the level of blood components would vary in patients in the course of the disease. Therefore, we considered four observation periods (day 1, 3, 7, and 14) for each patient in this course of COVID-19 infection. We collected blood samples from these patients on respective days and measured the level of blood hemoglobin (Hb), red blood cell (RBC), white blood cell (WBC), platelet, neutrophil, and blood oxygen (SPO2).

On day 1, we observed a downregulated Hb level (approximately 11.5 g/dL; SEM: 0.4008) among the female COVID-19 patients, where the normal range of Hb level is 12.1-15.1 g/dL. The Hb level among them was then found to be gradually increased to the normal level by day 3 with a slight decrease to approximately 12.1 (SEM: 0.621) on day 7. In regard to the male COVID-19 patients, the Hb level goes down to approximately 13.6 g/dL (SEM: 0.3602) from 14.0 g/dL on day 7 and then went up to the normal level (13.8-17.2 g/dL) again on day 14 (**Figure 3-A,B**). This result suggests that blood Hb level is slightly downregulated among male and female COVID-19 patients in the early days of disease progression.

**Figure 3:**
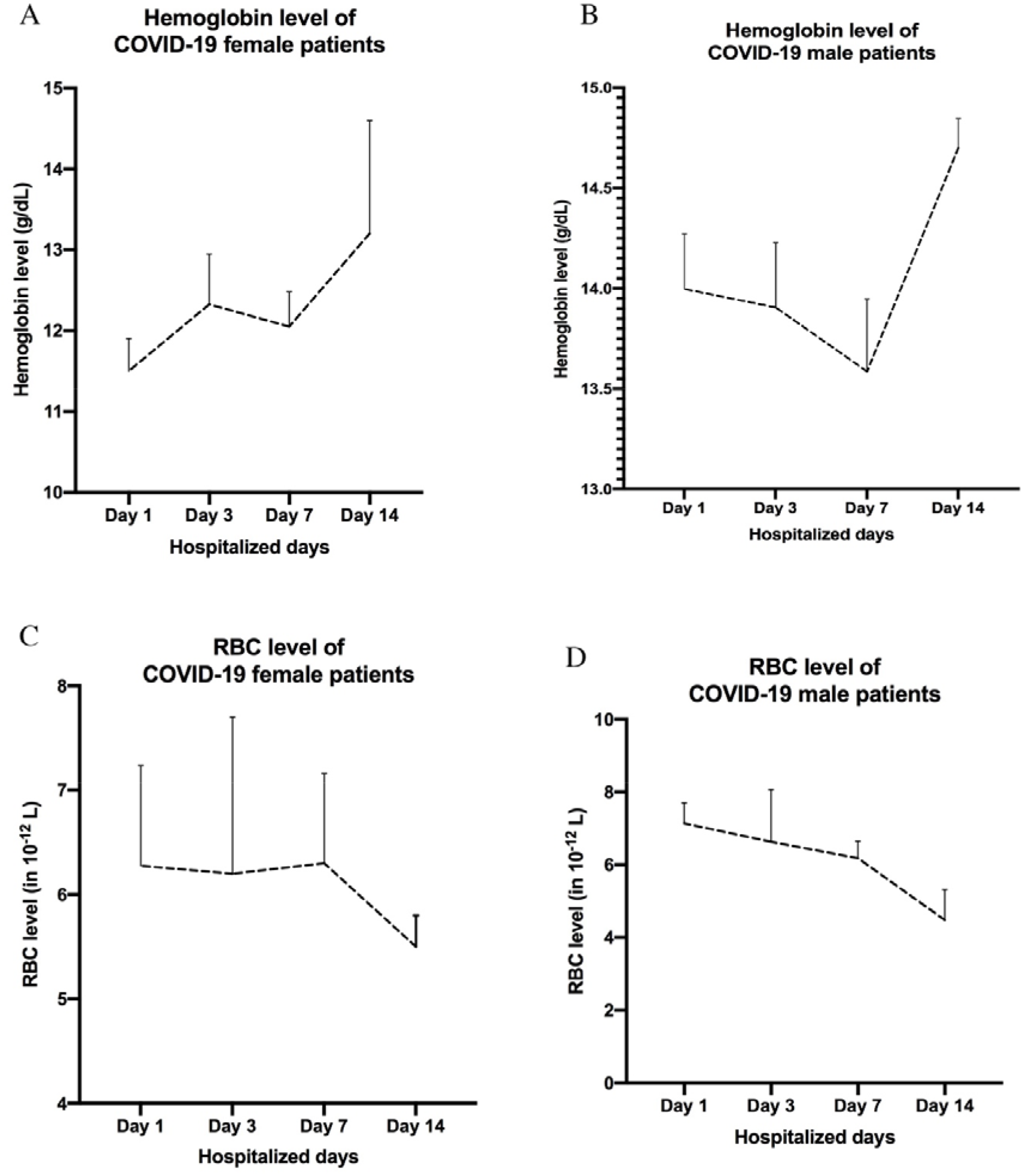
Determination of the level of blood hemoglobin and red blood cells in COVID-19 patients. Hemoglobin level was analyzed in both female (A) and male (B) patients. RBC level was also analyzed in both female (C) and male (D) patients. The graph was prepared using GraphPad Prism v9 software package. The represented data denotes Mean±SEM.

We observed an increased level of RBC from day 1 to day 7 in both male (normal range is 4.5-5.9⨯10^12^/L) and female (normal range is 4.1-5.1⨯10^12^/L) COVID-19 patients compared to the normal range as shown in **Figure 3-C,D**. However, after 7 days of disease symptoms, RBC level went down to the normal range.

Infection with CoV-2019 may not alter the level of SPO2, neutrophil, blood platelet, and WBC. However, the level of blood platelet and WBC were observed gradually upregulated within the normal range from day 3 to day 14 (data not shown).

### Level of serum components in COVID-19 patients

As the level of some of the blood components was differentially influenced by COVID-19, we tried to analyze the effect of this disease on the level of serum components in the hospital-admitted patients during the course of the disease. Therefore, we measured the level of serum albumin, bilirubin, creatinine, sodium, potassium, and chloride from the prepared serum samples of the COVID-19 patients.

We observed the average serum chloride level is downregulated to approximately 95 mEq/L (SEM: 0.7997) below the normal range (96-106 mEq/L) on day 14. However, infection with CoV-2019 does not significantly affect the level of serum albumin, bilirubin, creatinine, sodium, and potassium (supplementary **Figure S1**).

### High and sustained serum ALT level in COVID-19 patients

After observing the effect of COVID-19 on blood and serum components of the CoV-2019-infected patients, we hypothesized that COVID-19 influences the functionality of other organs of COVID-19 patients. Therefore, we analyzed the level of ALT/SGPT, AST/SGOT, and GCS scores in these patients during the course of the disease. ALT/SGPT is an enzyme, which is synthesized mainly by the liver, whereas the AST/SGOT is synthesized by multiple organs, including the liver, heart, etc.

Importantly, we observed a substantial increase in the level of ALT/SGPT (approximately 70 units/L; SEM: 13.56; normal range is 7-56 units/L) in COVID-19 patients on day 7 and was increased further to approximately 106 units/L (SEM: 12.64) on day 14. Likewise, we observed a significant increase in the level of AST/SGOT in COVID-19 patients even from day 1 (approximately 64.35 units/L; SEM: 5.013; normal range is 5-40 units/L). The level of AST/SGOT remained high in the duration of our 14-day observations and fluctuated, as shown in **Figure 4**. These results indicate that infection with CoV-2019 may affect the functionality of other organs of COVID-19 patients. Regarding the GCS score, only one patient was observed to present a score of 11 on day 4, and the patient sadly died on that day.

**Figure 4:**
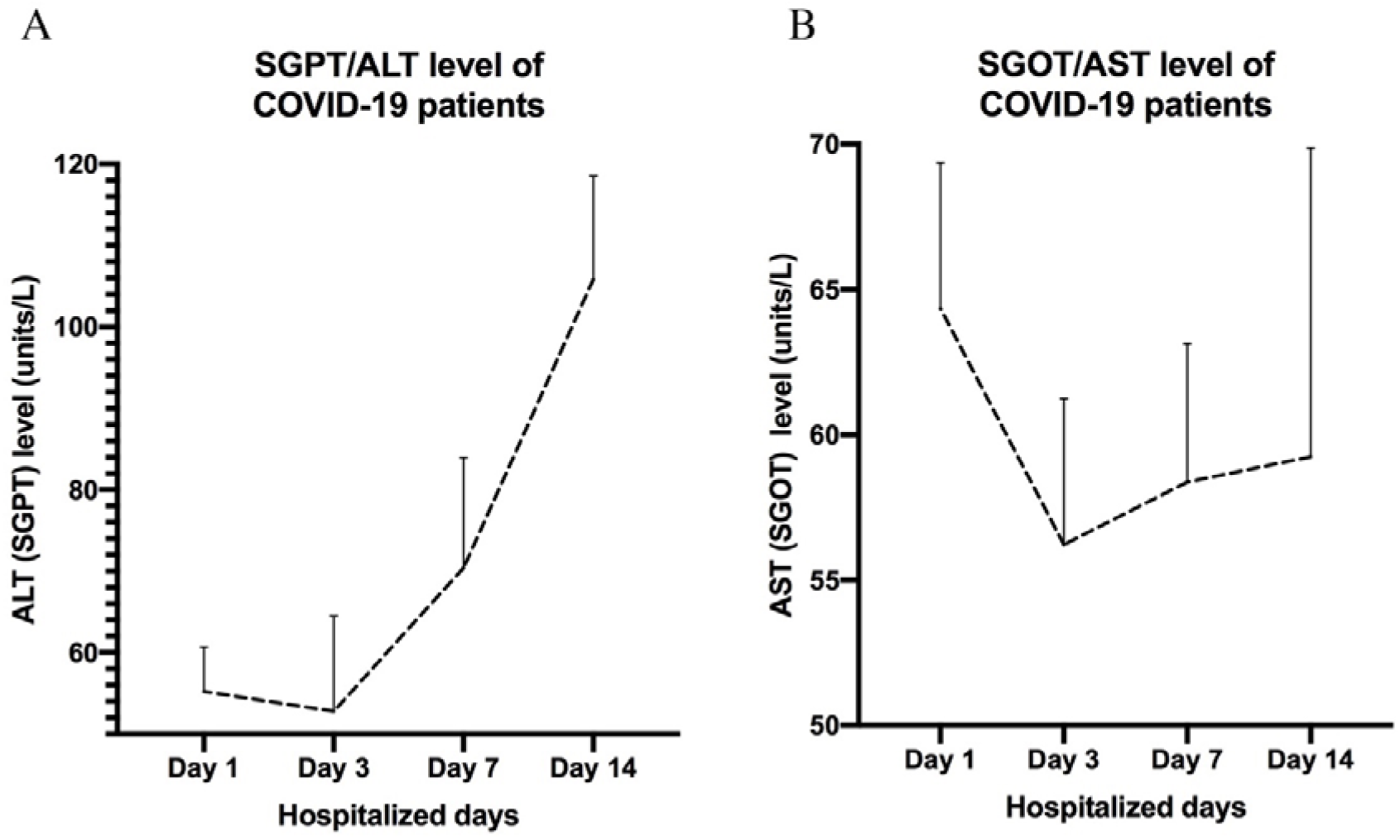
Analysis of the ALT (A) and AST (B) level in the serum of COVID-19 patients. The graph was prepared using GraphPad Prism v9 software package. The represented data denotes Mean±SEM

## Discussion

The severity of COVID-19 exceeds other respiratory viral infections regarding infectiousness as well as global mortality. Although there has been a tremendous increase in COVID-19 research since the commencement of the pandemic, some major research gaps still need to be addressed to better understand the nature of infection and the subsequent diseases contributed by CoV-2019. One of the critical research gaps seems to be the observation of the infected patients’ daily clinical manifestations. Therefore, in this study, we analyzed the clinical progression of the COVID-19 disease through a regular observation of biological parameters among the hospital-admitted COVID-19 patients over 14 days.

On day 1, approximately 19% of the patients developed pneumonia. Also, the overall respiratory rate of the patients was higher than the normal range on day 1, which is consistent with other studies with COVID-19 patients (Zhou et al. 2020). The increase in respiratory rate was observed heightened during the course of disease till day 14. Though the patients were not tested for chest X-ray in the subsequent days, the gradual increase in the respiratory rates may suggest that the disease was still influencing the patient’s lung over time.

Our findings noted hypertension and diabetes were the most common comorbid condition among the cases admitted in hospitals in Bangladesh and elsewhere (M.J. Hasan, Anam, et al. 2021; I. Hossain et al. 2020; Mowla et al. 2020). This finding reaffirmed observations reported in previous studies. One study with a sample of 100 admitted cases in ‘Dhaka Medical College Hospital, the largest teaching hospital in Bangladesh, reported the prevalence as hypertension (21%), diabetes mellitus (16%), ischemic heart disease (8%) among the admitted cases (Mowla et al. 2020). However, a single-center retrospective study with a sample of 157 hospitalized patients reported diabetes mellitus (24.8%) as the highest comorbid condition followed by hypertension (23.2%). This study’s inclusion criteria might be explained the relatively higher prevalence of diabetes mellitus, as this study included patients with at least one comorbid condition (M.J. Hasan, Anam, et al. 2021). Another study with a nearly similar sample size (n=33) reported diabetes mellitus (15%) and hypertension (27%) with the hospitalized patients (Bhuyan et al. 2020). Studies conducted in similar settings reported hypertension and diabetes as the commonly comorbid conditions among the hospitalized patients; for example, a nationwide analysis in China (Guan et al. 2020) reported hypertension (19.9%), diabetes (8.2%). Additional to the single study, systematic reviews (Ng et al. 2021; Sanyaolu et al. 2020) and meta-analysis reported hypertension and diabetes as the common comorbid conditions among people infected by the virus.

The COVID-19 patients were observed to have a reduced Hb level than the normal range on day 1, which is aligned with a study conducted with COVID-19 ICU patients (Lippi and Mattiuzzi 2020). This is a unique feature of CoV-2019 infection, which may indicate that the virus is acquiring the iron from the patient’s body for their survival and replication, though more research is needed to confirm this. Hb is essential to transfer oxygen from the lung to the tissues through the blood. Interestingly, though there is a lower Hb level in the patients’ blood, the level of oxygen was not altered enough.

COVID-19 was observed to influence the functionality of other body organs. Serum ALT/SGPT level of the patients was found enormously heightened after 7 days of disease progression than the normal range and sustained upregulated till day-14. Serum ALT is a potential marker of liver functionality (Gowda et al. 2009). The normal range of this enzyme is 7-56 units per liter of serum. On day 14 of our observation, we found that the level of ALT went significantly up by more than twice (108 units/L) than the normal range. Likewise, the level of AST was observed strongly high (approximately 64 units/L) on day 1 among the COVID-19 patients than its normal range 5-40 units/L and remained heightened during the course of observation. A high level of ALT and AST indicates the abnormality of liver function in the patients’ bodies, and it provokes a risk of liver damage (Gowda et al. 2009). Heightened AST level further denotes a risk of heart failure. Therefore, these results suggest that infection with CoV-2019 poses threat to not only the respiratory tract organ, but also to the liver as well as the heart.

In this study, we found one patient who sadly died of COVID-19 on day 4 after hospital admission. That patient was in 56-60 age range who had the comorbidity of hypertension and presented symptoms of fever and breathing difficulties from day 1 of hospital admission. The respiratory rate was consistently high (average of 42.4 breaths/min) and slightly low blood pressure (110/70 on day 3 and 80/50 on day 4). The GCS rate was regular (GCS 15) in the first two days of observation. However, on day 3 and 4, GCS rate went much lower to 12 and 11, respectively. Notably, the patient had already developed pneumonia on day 1 of hospital admission.

## Conclusions

This study presents a novel 14-day clinical observation of COVID-19 patients and is significant for three main reasons. Firstly, we present the potential comorbidities and wide ranges of symptoms shown by the COVID-19 patients. Secondly, the COVID-19 patients were essentially in our observation for approximately 14 days, and the clinical manifestations during the observations were analyzed. Thirdly and most importantly, the association of COVID-19 with subsequent liver and heart functionalities was identified by assessing the level of ALT and AST. We believe that the data of this study will help clinicians to decide the appropriate management of COVID-19 patients and take necessary actions to prevent the failure of other organs of the affected patients.

## Data Availability

Data is described through out the manuscript

## Data Availability

Data is decribed throughout the manuscript.

## Aknowledgement

All authors of this manuscript show highest gratitude to the participants of this study.

## Supplementary Figure Legends

**Supplementary Figure S1**: A 14-day observation of the level of serum components chloride (A), sodium (B), potassium (C), creatinine (D), albumin (E), and bilirubin (F) synthesized in COVID-19 patients. The graph was prepared using GraphPad Prism v9 software package. The represented data denotes Mean±SEM.

