## Supplementary figure s1 for "Clinical manifestations of hospitalized COVID-19 patients in Bangladesh: a 14-day observational study"

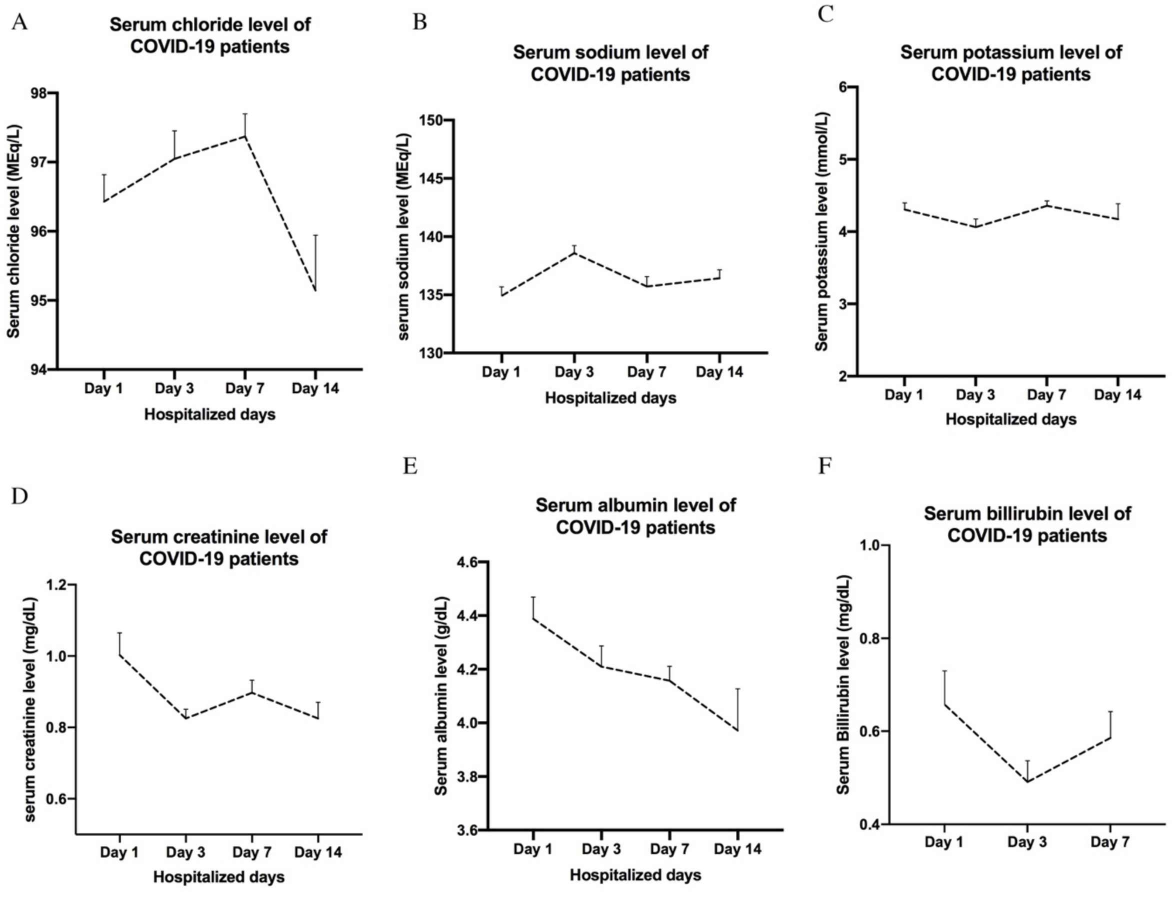


**Supplementary Figure S1**: A 14-day observation of the level of serum components chloride (A), sodium (B), potassium (C), creatinine (D), albumin (E), and bilirubin (F) synthesized in COVID-19 patients. The graph was prepared using GraphPad Prism v9 software package. The represented data denotes Mean±SEM.
